# Monosodium Urate Crystals within Cardiomyocytes

**DOI:** 10.64898/2026.01.20.26344247

**Authors:** Jiangwei Ma, Jing Tan, Conghui Shen, Tingting Mai, Ludong Yuan, Teng Wu, Shijie Xiong, Tongsheng Huang, Yuanjun Ji, Mengying Liu, Maoquan Yang, Erwen Huang, Weibin Cai

## Abstract

**Background:** While monosodium urate (MSU) crystal deposition within coronary plaques links hyperuricemia/gout to cardiovascular risk, the existence and functional consequences of MSU deposits within the myocardial interstitium or cardiomyocytes remain unknown.

**Methods:** Forensic autopsy myocardial and renal specimens from gout patients (n=4) and controls (n=4) were analyzed for MSU crystals using compensated polarized microscopy, laser capture microdissection coupled with HPLC-MS, and cryo-electron microscopy (cryo-EM) with electron diffraction. Pharmacological (adenine-diet) and genetic (Uox-/-) hyperuricemia murine models were assessed by crystal detection, echocardiography, and correlative histopathology.

**Results:** Myocardial MSU crystals were identified in all human gout cases (absent in controls), localized perivascularly, interstitially, and intracellularly within cardiomyocytes. Definitive verification was provided by compositional analysis (HPLC-MS m/z 167) and crystal structure confirmation (cryo-EM diffraction matching MSU). Hyperuricemic mice exhibited myocardial MSU deposition, progressive diastolic dysfunction (r =0.7014 vs. crystal burden), and reduced survival associated with higher crystal load. Importantly, myocardial crystal burden correlated more strongly with cardiac impairment than serum uric acid levels.

**Conclusion:** This study provides conclusive morphological, chemical, and crystallographic evidence of intracardiac MSU deposition in gout. It directly links myocardial crystal accumulation to diastolic dysfunction, proposing “uric acid cardiomyopathy” as a novel disease entity. Collectively, these findings achieve a paradigm shift in the perception of gout, redefining it from a peripheral arthropathy to a systemic disorder capable of causing direct myocardial injury.

## MAIN TEXT

Growing evidence confirms the presence of monosodium urate (MSU) crystals within coronary artery plaques in gout patients, solidifying the association between hyperuricemia/gout and heightened cardiovascular risk^1-3^. While dual-energy computed tomography (DECT) can detect relatively large ≥2mm MSU deposits^4^, the potential impact of microcrystalline MSU within the myocardium itself—either residing in the interstitium or intracellularly within cardiomyocytes—remains largely unexplored. Critically, within the milieu of a continuously contracting heart, even micrometer-sized MSU crystals, if present, could inflict significant structural and functional damage. To date, the existence or formation of MSU crystals directly within the myocardial tissue or cardiomyocytes has not been substantiated.

We examined cardiac and renal tissues from both patients with a history of gout and hyperuricemia mouse models, utilizing three experimental approaches to collectively verify the presence of MSU crystals within myocardial tissue. Human subject research was approved by the Ethics Committee of Sun Yat-sen University (No. 2024-062) and informed consents were obtained from the bereaved of victims. Animal experiments were conducted under protocols approved by the Institutional Animal Care and Use Committee of Sun Yat-sen University (SYSU-IACUC-2021-000857) (Figure [A]). In forensic autopsy cohort study, myocardial and renal specimens were examined from eight decedents, including four individuals with documented gout history (mean [SD] age, healthy group: 49.5 [7.8] years; hyperuricemia group: 46.3 [16.2] years). Given the established predisposition for renal MSU deposition in gout, renal sections were initially assessed via compensated polarized light microscopy to confirm crystal presence, serving as a positive internal control. Notably, identical analytical methodology revealed widespread MSU crystal deposition within myocardial tissues of all four gout cases (Figure [B-C]). Crystal distribution was systematically quantified across myocardial compartments—categorizing locations (perivascular, interstitial, intracellular cardiomyocytes) and crystal burden (dim, sparse, multiple small crystals, >3-μm aggregates). Analysis demonstrated the absence of detectable myocardial crystals in non-gout cases, contrasting with ubiquitous deposition in gout specimens. Furthermore, crystal aggregates were identified within perivascular spaces, interstitium, and cardiomyocyte interiors across all gout samples, with Case 4 exhibiting exceptionally severe deposition suggestive of potential clinical-pathological correlation (Figure [D]).

**Figure.**
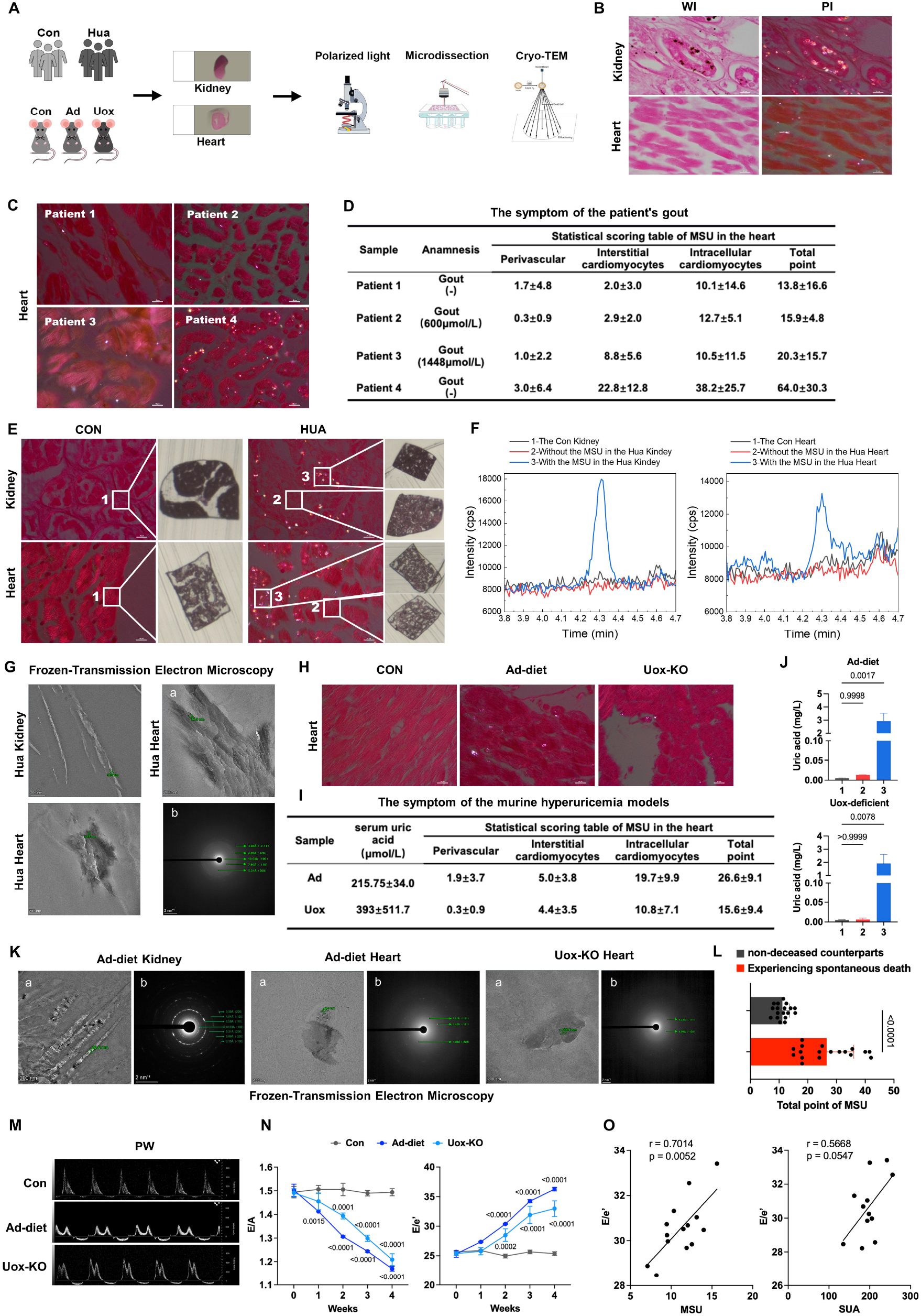
myocardial MSU crystal deposition exhibits the association with impaired cardiac function. **A**, Schematic of the strategy to identify the MSU with the forensic autopsy cohort (Con: healthy group; Hua: hyperuricemia group) and the adenine-induced model (0.25% adenine diet) and urate oxidase (Uox) knockout model. **B**, Kidney and heart sections from normal and hyperuricemic forensic samples under white light (WI) and polarized light (PI). PI: the presence of pathognomonic, birefringent MSU crystals. Scale bar, 10 µm. **C**, Heart sections from four hyperuricemic forensic samples under PI. **D**, Anamnesis based on *Forensic Appraisal Report*, (−) means serum uric acid not shown. Quantitative analysis of MSU deposition: morphometric analysis was performed on four hyperuricemic forensic samples. For each case, 20 randomly selected fields of view were analyzed to quantify MSU crystal locations and crystal burden (dim (0.5), sparse (1), multiple small crystals (2), >3-μm aggregates (4)). The table summarizes the results, presenting data as mean ± SD. **E**, LCM procurement of specific tissue regions under PI guidance. PI images show the precise excision of identically sized areas from: (1) normal tissue, (2) hyperuricemic tissue devoid of crystals, and (3) hyperuricemic tissue containing MSU crystals (bright birefringent deposits). The collected samples were analyzed by mass spectrometry. **F**, Quantitative MS analysis of uric acid in targeted tissues collected via LCM. **G**, Cryo-electron microscopy and crystallographic identification of MSU crystals. The corresponding selected-area electron diffraction (SAED) pattern obtained from the crystal in cryo-EM image. Indexing of the diffraction spots confirms a monoclinic crystal lattice with unit cell parameters matching the reference data for monosodium urate monohydrate. Scale bars: a, 200 nm; b, 2 nm?^1^. All the diffraction peaks can be indexed to the triclinic unit cell of MSU crystals with a=10.888, b=9.534, c=3.567Å, α=95.06°, β=99.47°, and γ=97.17°, space group P [1 with combining macron]. **H**, heart sections from normal and hyperuricemic mouse models under PI. **I**, Quantitative analysis of MSU deposition on two hyperuricemic mouse models. **J**, LCM procurement of specific tissue regions under PI guidance on two hyperuricemic mouse models and quantitative MS analysis of uric acid in targeted tissues collected via LCM. Bar graph showing the relative abundance of uric acid from three groups (1. The Con Heart; 2. Without the MSU in the Hua Heart; 3. With the MSU in the Hua Heart). Data are presented as mean ± SEM. Statistical significance was determined by one-way ANOVA followed by Tukey’s post-hoc test. **K**, Cryo-electron microscopy and crystallographic identification of MSU crystals. **L**, Bar graph showing the MSU deposition in mice experiencing spontaneous death before week 5 and non-deceased counterparts. Data are presented as mean ± SEM. Statistical significance was determined by unpaired Student’s t-test (two-tailed). **M-N**, Assessment of cardiac diastolic function (ratio between mitral E wave and A wave [E/A], and ratio between mitral E wave and E′ wave [E/E′]) in mice at week 4 after Ad-diet. Data are presented as mean ± SEM. Statistical significance was determined by two-way ANOVA test with Bonferroni multiple comparisons test. **O**, Correlation of cardiac function with pathological parameters. Scatter plots with linear regression lines depict significant correlations between echocardiographic measurement of cardiac function with the histologically quantified degree of MSU crystal deposition and level of serum uric acid. Statistical significance was assessed using Pearson/Spearman correlation analysis (r = correlation coefficient, p value shown).

To definitively ascertain whether the birefringent crystals observed under polarized light represented MSU, laser capture microdissection (LCM) was employed to precisely isolate these specific crystalline regions from tissue sections for unambiguous compositional analysis. Concurrently, adjacent non-birefringent myocardial tissue areas from gout samples and myocardial tissues from non-gout decedents were isolated as negative controls (Figure [E]). Microdissected samples underwent targeted analysis via high-performance liquid chromatography-mass spectrometry (HPLC-MS). This targeted approach successfully detected a dominant signal, the mass-to-charge ratio (m/z 167) and retention time (4.293 min) of which were identical to a purified MSU standard (Figure [F]). The direct correlation between the morphologically identified birefringent crystals and definitive chemical identification by MS provides conclusive evidence for the presence of MSU within the myocardial tissues of gout patients.

Direct cryo-electron microscopic examination of tissue specimens revealed crystalline deposits exhibiting marked acicular habit (30–50 nm thick) consistent with MSU. To obtain definitive crystallographic verification, individual crystals were subjected to electron diffraction analysis. The resultant diffraction patterns generated a unique set of lattice spacing (d-spacing) values and interplanar angles exclusively characteristic of MSU. These experimentally derived crystallographic parameters demonstrated unequivocal concordance with established reference data for synthetic MSU crystals, thereby definitively confirming the presence of pathologically significant MSU crystals within the myocardial tissue microenvironment under investigation (Figure [G]).

Demonstrating intracellular urate crystallization in cardiomyocytes questions the previously assumed resistance of the myocardial microenvironment (high flow, neutral pH, local urate concentration). Convergent evidence from integrated morphological, chemical, and crystallographic analyses provides definitive verification of myocardial MSU crystal deposition. To investigate whether MSU deposition exacerbates myocardial injury and cardiac dysfunction, two mouse models of hyperuricemia—pharmacologically induced and genetic urate oxidase (Uox) knockout—were employed (Figure [H]).

Polarized light microscopy, mass spectrometric identification, andcryo-electron microscopy collectively confirmed the presence of MSU crystals within myocardial tissues of both adenine-diet and Uox-deficient mice (Figure [I-K]). Serial echocardiographic assessment in hyperuricemic mice revealed progressive impairment of diastolic function emerging at four weeks post-induction, while systolic contractile function remained initially preserved (Figure [M-N]). Notably, the severity of diastolic dysfunction correlated with more myocardial MSU deposition, suggesting that hyperuricemia and gout may accelerate cardiac functional decline through intracardiac MSU crystallization (Figure [O]). Notably, Intramyocardial MSU crystal deposition correlates more strongly with diastolic dysfunction severity than serum uric acid levels (r=0.7014 *vs* 0.5668). Meanwhile, we observed that under the adenine-diet, mice experiencing spontaneous death before week 5 exhibited significantly greater myocardial MSU crystal deposition than non-deceased counterparts (Figure [J]). Combined with forensic autopsy cohort study, these findings collectively indicate that serum uric acid levels in gout patients do not necessarily totally reflect the extent of MSU crystal deposition within the myocardium. Furthermore, myocardial MSU deposition may serve as a stronger predictor of cardiac functional impairment and lifespan.

In summary, this study provides reliable evidence from human hyperuricemia/gout and corroborative murine models demonstrating the definitive deposition of MSU crystals within myocardial tissue—localized to both interstitial and intracellular compartments. This crystallopathy exhibits a sgnificant association with the impaired diastolic function. It may define a new type of heart disease (“uric acid cardiomyopathy” or “cardiac gout”)—that elucidates the pathogenesis of previously unexplained arrhythmogenic and cardiomyopathic states. The robust association between intracardiac MSU crystallization and functional impairment underscores the imperative for elevating serum urate and MSU deposition monitoring to a cardiovascular risk stratification parameter commensurate with cholesterol and glucose assessment. This work fundamentally redefines the systemic repercussions of gout, transforming its clinical perception from a peripheral arthropathy to a disorder conferring significant direct cardiac morbidity.

## Data Availability

All data produced in the present study are available upon reasonable request to the authors

## Acknowledgments

We gratefully acknowledge Dr. Shuyao Huang and Ms. Hui Qin of Instrumental Analysis & Research Center, Prof. Qinfen Zhang and Dr. Hongmei Li of the State Key Laboratory of Biocontrol, Sun Yat-sen University, for their great help and technical support in the Cryo-electron microscopy and crystallographic identification of MSU crystals.

